# Accelerating the pace and accuracy of systematic reviews using AI: a validation study

**DOI:** 10.1101/2024.12.10.24318803

**Authors:** Jiada Zhan, Kara Suvada, Muwu Xu, Wenya Tian, Kelly C. Cara, Taylor C. Wallace, Mohammed K. Ali

**Author notes:** Corresponding author: Jiada Zhan; address: 1518 Clifton Road, Atlanta, Georgia 30322 USA Kara Suvada; address: 1518 Clifton Road, Atlanta, Georgia 30322 USA). Co-first authors (Jiada Zhan, Kara Suvada).

## Abstract

**Background:** Artificial intelligence (AI) can greatly enhance efficiency in systematic literature reviews and meta-analyses, but its accuracy in screening titles/abstracts and full-text articles is uncertain.

**Objectives:** This study evaluated the performance metrics (sensitivity, specificity) of a GPT-4 AI program, Review Copilot, against human decisions (gold standard) in screening titles/abstracts and full-text articles from four published systematic reviews/meta-analyses.

**Research Design:** Participant data from four already-published systematic literature reviews were used for this validation study. This was a study comparing Review Copilot to human decision-making (gold standard) in screening titles/abstracts and full-text articles for systematic reviews/meta-analyses. The four studies that were used in this study included observational studies and randomized control trials. Review Copilot operates on the OpenAI, GPT-4 server. We examined the performance metrics of Review Copilot to include and exclude titles/abstracts and full-text articles as compared to human decisions in four systematic reviews/meta-analyses. Sensitivity, specificity, and balanced accuracy of title/abstract and full-text screening were compared between Review Copilot and human decisions.

**Results:** Review Copilot’s sensitivity and specificity for title/abstract screening were 99.2% and 83.6%, respectively, and 97.6% and 47.4% for full-text screening. The average agreement between two runs was 95.4%, with a kappa statistic of 0.83. Review Copilot screened in one-quarter of the time compared to humans.

**Conclusions:** AI use in systematic reviews and meta-analyses is inevitable. Health researchers must understand these technologies’ strengths and limitations to ethically leverage them for research efficiency and evidence-based decision-making in health.

## Background

The landscape of scientific research is growing profoundly with an exponential number of scholarly manuscripts being added to online and print media on a wide array of topics.^1^ This proliferation of scientific data offers immense opportunities for knowledge acquisition and dissemination but simultaneously presents formidable challenges in information management and synthesis. Systematic reviews and meta-analyses are crucial methodologies for synthesizing this vast body of literature by offering comprehensive, replicable, and transparent overviews of various subjects.^2^ They are critical to researchers, clinicians, guideline committees, decision-makers, and the general public.

Despite their critical importance, systematic reviews remain labor-intensive (i.e., 1,000+ hours) and are often constrained by the necessity of narrow search strategies that balance the recall and precision of information retrieval.^3,4^ The challenge is compounded by the rapid growth of literature in many fields that are important to the topic, making comprehensive, manual review increasingly unfeasible.^5^ The traditional workflow of a systematic review includes three main time-consuming components: abstract and title screening, full-text screening, and data extraction.

Machine learning offers opportunities for assisting researchers in completing systematic reviews during the title and abstract screening stage. The most popular machine learning technique is active learning, which is a subset of machine learning that involves a semi-automated approach where the algorithm actively queries the user to label data points and shuffle the order of all records. This allows active learning to present the most relevant articles recommended by the algorithm to researchers. There are many existing active learning tools, including Abstrackr,^6^ ASReview,^7^ Colandr,^8^ FASTREAD,^9^ Rayyan,^10^ RobotAnalyst^11^, Covidence^12^, and DistillerSR^13^. However, existing active learning tools have three major limitations. First, to our knowledge, existing tools often lack the versatility needed to screen articles with a diverse range of research fields. Second, validation is mostly lacking in details in many machine learning tools for systematic reviews.^7^ Third, active learning tools define stopping rules in arbitrary and uncertain ways. The stopping rule is when a model predicts none of the remaining abstracts to be relevant. Usually, an arbitrary cutoff such as 100 irrelevant articles screened in a sequence is used. Theoretically, reviewing all abstracts using alternative approaches is more reliable than selecting a specific stopping rule.

To address these limitations, we developed “Review Copilot,” an innovative, artificial intelligence (AI) application (a large language model pipeline) designed specifically for accelerating systematic reviews. Review Copilot uses GPT-4^14^ to address the aforementioned gaps by offering flexibility and the ability to handle a wide array of research topics automatically under the supervision of researchers. Its benchmark testing also allows for the critical evaluation of algorithm performance in real-world scenarios. In this study, we tested Review Copilot decisions against human decisions using four already-published, human-reviewed systematic reviews/meta-analyses. We reported the performance (i.e., sensitivity and specificity) of Review Copilot for title and abstract screening, full-text screening, and overall screening performance. All manuscript text is reported using STROBE guidelines for reporting in this manuscript (**Supplemental Digital Content: Table 1**).^15^

**Table 1.** Description of studies used for Review Copilot performance metrics and validation.

| Citation | Systematic literature review, meta-analysis, or both? | Type of studies included in SLR/MA | Types of decisions reviewed | Total number of person hour spent per researcher | Total number of person hour spent by AI |
| --- | --- | --- | --- | --- | --- |
| Cara KC, Ar B, Tc W, M C. Safety of Using Enteral Nutrition Formulations Containing Dietary Fiber in Hospitalized Critical Care Patients: A Systematic Review and Meta-Analysis. *JPEN J Parenter Enteral Nutr.* 2021;45(5). doi:10.1002/jpen.2210 | Both | Almost all study types, except narrative reviews, editor letters, protocol, background reading, and abstracts | Title/abstract exclusion; full-text screening; full-text inclusion | 10-12 hours | 3.5 |
| Cara KC, Beauchesne AR, Wallace TC, Chung M. Effects of 100% Orange Juice on Markers of Inflammation and Oxidation in Healthy and At-Risk Adult Populations: A Scoping Review, Systematic Review, and Meta-analysis. *Adv Nutr.* 2021;13(1):116-137. doi:10.1093/advances/nmab101 | Both | Any intervention and observational studies | Title/abstract exclusion; full-text screening; full-text inclusion | 10-12 hours | 6.5 |
| Galaviz KI, Weber MB, Suvada K, et al. Interventions for Reversing Prediabetes: A Systematic Review and Meta-Analysis. *Am J Prev Med.* 2022;62(4):614-625. doi:10.1016/j.amepre.2021.10.020 | Both | Randomized control trials | Full-text inclusion; general exclusion | 40-50 hours | 13 |
| Meijboom RW, Gardarsdottir H, Egberts TCG, Giezen TJ. Patients Retransitioning from Biosimilar TNFα Inhibitor to the Corresponding Originator After Initial Transitioning to the Biosimilar: A Systematic Review. *BioDrugs Clin Immunother Biopharm Gene Ther.* 2022;36(1):27-39. doi:10.1007/s40259-021-00508-4 | Systematic literature review | Randomized control trials and observational studies | Full-text inclusion; general exclusion | 75-90 hours | 4.5 |

## Methods & Measures

We describe both the pipeline development and validation approach.

### Pipeline description

We used the latest large language model from OpenAI, GPT-4, to develop Review Copilot. Review Copilot follows the population, intervention, comparison, outcome, and study type (PICOS) framework while reviewing relevant information in databases and making decisions. Prompt engineering was performed to ensure Review Copilot will always follow the instruction of the PICOS with a consistent format in different scenarios.

For abstract and title screening and full text screening, Review Copilot requires researchers to first define their research goals using the PICOS framework.^3^ Then, researchers can provide the list of titles and abstracts to be screened via CSV format or the full text of articles to be screened via PDF format to Review Copilot. Subsequently, Review Copilot can extract relevant information from the articles and make decisions by comparing the extracted PICOS with the target PICOS. Decisions made by Review Copilot are accompanied by rationales, such as recognizing the wrong population, wrong exposure, wrong outcomes, etc. The researcher can then make final decisions based on the AI’s decisions and justifications.

### Description of validation datasets

We used four already-published systematic reviews as datasets for this validation study. The first validation dataset (Cara et al., 2021 Fiber) was from a systematic review and meta-analysis that included articles with almost all study types, except narrative reviews, editor letters, protocols, background reading, and abstracts, regarding the safety of using enteral formula with dietary fiber in hospitalized critical care patients.^16^ After removing duplicates, 482 articles from search databases were included in the title and abstract screening stage. Among these articles, based on human reviews, 429 articles were excluded during title/subtract screening, and 34 articles were excluded during the full-text screening; 19 articles were finally included in the review.

The second validation dataset (Cara et al., 2021 Juice) was from a systematic review and meta-analysis of any intervention and observational studies assessing the associations between 100% orange juice and biomarkers of inflammation and oxidation in generally healthy populations.^17^ After removing duplicates, 1183 articles from search databases remained for title and abstract screening. Among these studies, in human review, 1098 articles were excluded during the title/subtract screening, and 63 articles were excluded during the full-text screening; 22 articles were included in the final review.

The third validation dataset (Galaviz et al., 2022) was from a systematic review and meta-analysis of randomized controlled trial (RCTs) evaluating interventions for reversing prediabetes in adults^18^. After deduplication, 3547 articles from literature databases were included for screening. Through human review, 3502 articles were excluded during the title/abstract and the full-text screening phases; 45 articles were included in the final review.

The fourth validation dataset (Meijboom et al., 2022) was from a systematic review of RCTs and observational studies related to the incidence of outcomes of patients with cancer who transitioned from one cancer drug to another versus patients who did not transition drugs.^19^ After deduplication, 994 articles from literature searches were included for screenings. In human review, 957 articles were excluded during the title/abstract and full-text screenings. Finally, 37 articles were included in the pooled analysis.

The estimated time it took to complete title/abstract screening and full-text screening together for Cara et al., 2021 (Fiber), Cara et al., 2021 (Juice), and Galaviz et al., 2022 was 10-12 hours, 10-12 hours, and 40-50 hours, respectively (Meijboom et al., 2022 have not responded to our request for the approximate number of human hours) (**Table 1**).

### Validation and performance metrics of Review Copilot

To validate and assess the performance of Review Copilot, we used human researcher-extracted data as the gold standard from four systematic reviews or meta-analyses that had already been published.^16–19^ We reviewed the PROSPERO protocols for all four studies and used the past human decisions related to title and abstract exclusion, full-text inclusion, full-text exclusion, and/or general exclusion.

We first assessed the performance of Review Copilot for each dataset for title/abstract and full-text screening. We also assessed the inter-rater reliability (kappa statistic) using two runs of the same AI pipeline for each dataset. For our supplementary analyses, we assessed the performance of the following: (1) title/abstract screening alone; (2) full-text screening alone.

The main performance metric was sensitivity, while the secondary performance metrics were specificity, balanced accuracy, and reliability. Sensitivity measures the proportion of true positives (inclusions according to the human gold standard) that are correctly identified by the AI. A high sensitivity in this context means that AI is effective at making a high proportion of the same decisions that a human expert would without missing many inclusions. Specificity assesses the proportion of true negatives (decisions where the human gold standard would indicate to exclude) that are correctly identified as such by the AI and indicates that the AI is proficient at avoiding false positives. Balanced accuracy is the sensitivity plus the specificity divided by two and is used as a secondary metric, and in our case, was used because of an imbalance between the number of human includes and excludes (there were many more exclusions as compared to inclusions). To assess the reliability, for every dataset, two runs were performed using the same AI pipeline. We evaluated the inter-rater reliability of two AI runs by percent agreement and kappa statistic.^20^ Sensitivity and specificity were evaluated using the *confusionMatrix* function in the *caret* package in R 4.2.2 (http://www.r-project.org/, The R Foundation, Vienna, Austria). The kappa statistic was calculated by using the *kappa2* function in the *irr* package in the same version of R.

### Inclusion and ethics statements

It was not possible for the authors of this study to involve the public in the design of this research study, as the datasets were provided by the first/senior authors of each of the four studies and a novel, GPT-4 application was used in the validation test. Additionally, the information of individuals who were included in these datasets were de-identified prior to using their data, so subject involvement would not be possible. With regards to dissemination, we plan to share results and recommendations with public health officials and clinicians through sharing our results widely throughout our professional networks via the publication of this manuscript. This study was a secondary data analysis and tested accuracy and validity of Review Copilot as compared to human decision-making. This work was determined non-human subjects research by the Emory University IRB.

## Results

The estimated time it took to Review Copilot to complete title/abstract screening and full-text screening together for Cara et al., 2021 (Fiber), Cara et al., 2021 (Juice), and Galaviz et al., 2022 and Meijboom et al., 2022 was 3.5 hours, 6.5 hours, 13 hours, and 4.5 hours, respectively (**Table 1**).

Sensitivity of Review Copilot for all four datasets for *title/abstract screening* was 99.2%. Overall specificity for title/abstract screening was 83.6%, and balanced accuracy was 91.4% (**Table 2**). When stratifying by each of the four datasets, Cara et al., 2021 (Fiber), Cara et al., 2021 (Juice), Galaviz et al., 2022, and Meijboom et al., 2022, title/abstract screening sensitivity and specificity were: 100.0% and 88.3%; 100.0% and 95.2%; 100.0% and 82.8%; and 97.3% and 69.2%, respectively (**Supplemental Digital Content: Tables 2 & 3**).

**Table 2.** The Performance of Review Copilot in Both Screening Stages of All Datasets.

| Screening stages | Average Performance | Overall |
| --- | --- | --- |
| Title and abstract screening | Sensitivity | 99.2% |
|  | Specificity | 83.6% |
|  | Balanced Accuracy | 91.4% |
|  | Percent Agreement | 95.4% |
|  | Kappa Statistic | 0.83 |
| Full-text screening | Sensitivity | 97.6% |
|  | Specificity | 47.4% |
|  | Balanced Accuracy | 72.5% |
|  | Percent Agreement | 89.8% |
|  | Kappa Statistic | 0.74 |

Sensitivity of Review Copilot for all four datasets for *full-text screening* was 97.6%. Overall specificity for full-text screening was 47.4% and balanced accuracy was 72.5% (**Table 2**). When stratifying by the two datasets with disaggregated full-text decisions, Cara et al., 2021 (Fiber) and Cara et al., 2021 (Juice), full-text specificity and sensitivity were: 100.0% and 52.4%; and 81.8% and 59.6%, respectively (**Supplemental Digital Content: Tables 2 & 3**).

We also constructed a confusion matrix with the number of true positives, false positives, false negatives, and true negatives classified by Review Copilot, as compared to the gold standard (human decisions), for all datasets in both title/abstract and full-text screening stages together There were 162 true positives, 992 false positives, 2 false negatives, and 5028 true negatives (**Figure 1**). The specific confusion matrix for each dataset for title/abstract screening and/or full-text screening is provided in the supplementary materials (**Supplemental Digital Content: Figures 1 & 2**).

**Figure 1:** Confusion Matrix for All Datasets. This figure depicts the concordance and non-concordance between the gold-standard, human decision for abstract/title and full-text inclusion/exclusion (x-axis) compared to the AI/Review Copilot decision (y-axis). For example, the AI included 162 articles that humans did, yet excluded 2 articles that humans included.

Regarding the reliability of Review Copilot in two separate runs in the title/abstract screening stage for all datasets, the average percent agreement between two Review Copilot’s runs was 95.4% and the average kappa statistic was 0.83. For the full-text screening stage, the average percent agreement was 89.8% and the average kappa statistic was 0.74 (**Table 2**).

## Discussion

At one-quarter of the time required to conduct reviews manually, the high sensitivity of Review Copilot shows the promise and potential of using AI technology for assisting researchers in speeding up, streamlining, and accurately screening literature during systematic reviews. High sensitivity indicates that the AI successfully included nearly all the articles that the human reviewers included; in fact, the AI only did not include one article each at both the title/abstract screening and full-text screening stage that the human did include. Lower specificity indicates that the AI included articles that the human excluded – this makes the AI less efficient at excluding articles that should have been excluded. An “over-inclusive” AI means the researcher would still need to review all the articles that the AI included before moving to the full-text screening stage but could be assured that the AI did not miss any articles that should have been included. Furthermore, human researchers would have to review significantly fewer articles following Review Copilot’s process than if they conducted a review without Review Copilot.

Within the past 18 months, the widespread uptake of ChatGPT has given rise to the question of using AI for mostly or fully automated systematic literature reviews/meta-analyses. Some studies have found that AI can make mostly accurate decisions regarding article inclusion, exclusion, and the identification of Boolean terms; many of these studies also found that AI, namely ChatGPT, can reduce the amount of time it takes for human researchers to conduct a systematic literature review/meta-analysis.^21–27^ One study found that initial title/abstract screening burden was reduced by up to 61%.^22^ None of these articles found that AI can fully automate the review process, however, citing pitfalls such as limited ability to extract full-text articles in real-time, researcher influence/bias on the algorithm used for article review, and/or uncharted ethical considerations.^21,27^ Additionally, thoughtful consideration needs to be placed on: 1) making sure AI applications do not include unethical research; and, 2) placing human understanding in the application and context of results.^26^

Our evaluation and validation of Review Copilot shows similar findings in that the tool can be used to streamline processes of the review in both the title and abstract screening and full-text screening stages, but it cannot entirely replace the work of the researcher. As Review Copilot can exclude about 90-95% of irrelevant articles during the title and abstract screening stage with the speed of 1000 articles per four person hours, even now, a researcher could potentially use Review Copilot to substantially reduce the amount of time it takes to include the required abstracts for the next stage of the review. The time saved screening abstracts and full-text articles could free up time for researchers to conduct additional analyses and communicate results more broadly. For all datasets, Review Copilot took approximately 66-75% fewer person-hours than humans when screening articles at both stages. Additionally, Review Copilot can also screen articles in the full-text stage and exclude about 50% of the irrelevant articles. The investigator would still have to review the remaining articles that the AI could not exclude, as the specificity of Review Copilot indicates that it does not yet have a high enough accuracy as to exclude all the necessary articles in both screening stages. As AI models improve even further, the accuracy of AI to discern the proper exclusions will surely also increase and could be used for other aspects of reviews, such as risk of bias/quality assessments and data extraction. This might be achieved by training the large language models on larger datasets.

Our study had key strengths. A major strength of our study is that we were able to compare Review Copilot results to peer-reviewed human decisions, for which authors generously shared their files for different stages of each review; all four systematic reviews that served as the datasets were already published and additionally went through rigorous internal validation among study authors. Our results are also generalizable to most study types and topics; the four systematic reviews/meta-analyses included both randomized controlled trials and observational studies across an array of topics (i.e., pediatric and adult nutrition, prediabetes interventions, and pharmaceutical oncology interventions). A limitation of this work was the smaller number of systematic reviews/meta-analyses used in our review (N=4). A higher number of reviewed studies would have increased the robustness of our results.

Using AI in systematic reviews dates to 2006, where neural networks were proposed to select primary studies based on text mining^27^. As we enter a more advanced era of AI technology, namely with the release of tools such as ChatGPT (OpenAI), Gemini (Google), Copilot (Microsoft), and ERNIE Bot (Baidu), the field of AI in systematic reviews is only expected to grow.^28^ Leveraging this technology can help generate evidence more efficiently, while simultaneously acknowledging the importance of human interpretation and application of results to health.^27^ No AI technology can successfully conduct a systematic review on its own in the current stage, including Review Copilot, indicating that researchers will still need methodological prowess and contextual knowledge to leverage AI for efficiency when conducting reviews and solving human problems.

## Conclusions

We tested a new AI application, Review Copilot, that uses GPT-4 to screen articles at the title/abstract screening and full-text screening stages of four already-published systematic literature reviews/meta-analyses against human decisions (gold standard). The AI’s sensitivity was high, while specificity was moderate. Therefore, it appears that Review Copilot could be a helpful and useful tool to streamline the screening processes for systematic reviews. More development and advancement of these tools is inevitable and warranted to improve our understanding of how to best leverage these technologies to advance scientific evidence in efficient, ethical, and methodologically-sound ways.

## Data Availability

The datasets used and/or analyzed during the current study are available from the corresponding author on reasonable request (email the corresponding authors at if interested).

## Code Availability

The underlying code for this study and validation datasets is not publicly available but may be made available to researchers on reasonable request from the corresponding authors.

## Prior presentations

None.

## Supplementary Digital Content: SupplementaryDigitalContent.pdf

Contains the following:

Supplemental Digital Content: Table 1: STROBE Statement—checklist of items that should be included in reports of observational studies.

Supplementary Digital Content: Table 2. Number of past human decisions of studies used for Review Copilot performance metrics and validation.

Supplementary Digital Content: Table 3. The performance of Review Copilot for all decisions in 4 datasets during the title/abstract screening and full-text screening stages.

Supplementary Digital Content: Figure 1. Confusion matrices of the four validation datasets (A-D) for the title and abstract screening.

*Figure legend: This figure depicts the concordance and non-concordance between the gold-standard, human decision for abstract/title inclusion/exclusion (x-axis) compared to the AI/Review Copilot decision (y-axis) for each dataset. A is Cara et al. (Fiber) (2021), B is Cara et al. (Juice) (2021), C is Galaviz et al. (2022), and D is Meijboom et al. (2022).*

Supplementary Digital Content: Figure 2. Confusion matrices of two of the datasets (A,B) for full-text screening

*Figure legend: This figure depicts the concordance and non-concordance between the gold-standard, human decision for full-text inclusion/exclusion (x-axis) compared to the AI/Review Copilot decision (y-axis) for each dataset. A is Cara et al. (Fiber) (2021), and B is Cara et al. (Juice) (2021). Datasets C (Galaviz et al. (2022)), and D (Meijboom et al. (2022)) could not be evaluated individually at the full-text level because of software restrictions used for these reviews.*

## References

1. Growth rates of modern science: A bibliometric analysis based on the number of publications and cited references - Bornmann - 2015 - Journal of the Association for Information Science and Technology - Wiley Online Library. https://asistdl.onlinelibrary.wiley.com/doi/abs/10.1002/asi.23329.

2. Uman, L. S. Systematic Reviews and Meta-Analyses. J Can Acad Child Adolesc Psychiatry 20, 57–59 (2011).

3. Higgins, J. P. & Green, S. Cochrane Handbook for Systematic Reviews of Interventions: Cochrane Book Series.

4. Allen, I. E. & Olkin, I. Estimating Time to Conduct a Meta-analysis From Number of Citations Retrieved. JAMA 282, 634–635 (1999).

5. Toward systematic review automation: a practical guide to using machine learning tools in research synthesis | Systematic Reviews. https://link.springer.com/article/10.1186/s13643-019-1074-9.

6. Wallace, B. C., Small, K., Brodley, C. E., Lau, J. & Trikalinos, T. A. Deploying an interactive machine learning system in an evidence-based practice center: abstrackr. in Proceedings of the 2nd ACM SIGHIT International Health Informatics Symposium. 819–824 (Association for Computing Machinery, New York, NY, USA, 2012). doi:10.1145/2110363.2110464.

7. van de Schoot, R. et al. An open source machine learning framework for efficient and transparent systematic reviews. Nat Mach Intell 3, 125–133 (2021).

8. Using machine learning to advance synthesis and use of conservation and environmental evidence - PubMed. https://pubmed.ncbi.nlm.nih.gov/29644722/.

9. [PDF] Finding better active learners for faster literature reviews | Semantic Scholar. https://www.semanticscholar.org/paper/Finding-better-active-learners-for-faster-reviews-Yu-Kraft/2e44e41490642365fd3d60e78c246b67e3bc7941.

10. Rayyan—a web and mobile app for systematic reviews | Systematic Reviews | Full Text. https://systematicreviewsjournal.biomedcentral.com/articles/10.1186/s13643-016-0384-4.

11. Przybyła, P. et al. Prioritising references for systematic reviews with RobotAnalyst: A user study. Res Synth Methods 9, 470–488 (2018).

12. Covidence - Better systematic review management. Covidence https://www.covidence.org/.

13. DistillerSR | Systematic Review Software | Literature Review Software. DistillerSR https://www.distillersr.com/products/distillersr-systematic-review-software.

14. ChatGPT. https://chat.openai.com.

15. von Elm, E. et al. The Strengthening the Reporting of Observational Studies in Epidemiology (STROBE) Statement: Guidelines for Reporting Observational Studies. Epidemiology 18, 800 (2007).

16. Kc, C., Ar, B., Tc, W. & M, C. Safety of Using Enteral Nutrition Formulations Containing Dietary Fiber in Hospitalized Critical Care Patients: A Systematic Review and Meta-Analysis. JPEN. Journal of parenteral and enteral nutrition 45, (2021).

17. Cara, K. C., Beauchesne, A. R., Wallace, T. C. & Chung, M. Effects of 100% Orange Juice on Markers of Inflammation and Oxidation in Healthy and At-Risk Adult Populations: A Scoping Review, Systematic Review, and Meta-analysis. Adv Nutr 13, 116–137 (2021).

18. Galaviz, K. I. et al. Interventions for Reversing Prediabetes: A Systematic Review and Meta-Analysis. American Journal of Preventive Medicine 62, 614–625 (2022).

19. Meijboom, R. W., Gardarsdottir, H., Egberts, T. C. G. & Giezen, T. J. Patients Retransitioning from Biosimilar TNFα Inhibitor to the Corresponding Originator After Initial Transitioning to the Biosimilar: A Systematic Review. BioDrugs 36, 27–39 (2022).

20. McHugh, M. L. Interrater reliability: the kappa statistic. Biochem Med (Zagreb) 22, 276–282 (2012).

21. Alshami, A., Elsayed, M., Ali, E., Eltoukhy, A. E. E. & Zayed, T. Harnessing the Power of ChatGPT for Automating Systematic Review Process: Methodology, Case Study, Limitations, and Future Directions. Systems 11, 351 (2023).

22. Kebede, M. M., Le Cornet, C. & Fortner, R. T. In-depth evaluation of machine learning methods for semi-automating article screening in a systematic review of mechanistic literature. Research Synthesis Methods 14, 156–172 (2023).

23. Blaizot, A. et al. Using artificial intelligence methods for systematic review in health sciences: A systematic review. Research Synthesis Methods 13, 353–362 (2022).

24. Khalil, H., Ameen, D. & Zarnegar, A. Tools to support the automation of systematic reviews: a scoping review. Journal of Clinical Epidemiology 144, 22–42 (2022).

25. Mahuli, S. A., Rai, A., Mahuli, A. V. & Kumar, A. Application ChatGPT in conducting systematic reviews and meta-analyses. British Dental Journal 235, 90–92 (2023).

26. Qureshi, R. et al. Are ChatGPT and large language models “the answer” to bringing us closer to systematic review automation? Systematic Reviews 12, 72 (2023).

27. Dijk, S. H. B. van et al. Artificial intelligence in systematic reviews: promising when appropriately used. BMJ Open 13, e072254 (2023).

28. de la Torre-López, J., Ramírez, A. & Romero, J. R. Artificial intelligence to automate the systematic review of scientific literature. Computing 105, 2171–2194 (2023).

